# Add-on PCSK9 Inhibitor Therapy for Acute Stroke Due to Intracranial Atherosclerosis: A Randomized Clinical Trial

**DOI:** 10.1101/2025.08.04.25332997

**Authors:** Yi-Ting Pan, Yuan-Hsiung Tsai, Hsu-Huei Weng, Jiann-Der Lee, Jen-Tsung Yang, Leng-Chieh Lin, Yen-Chu Huang

## Abstract

**Background:** Intracranial atherosclerotic stenosis (ICAS) is a major cause of ischemic stroke with high recurrence rates despite intensive therapy. The efficacy of proprotein convertase subtilisin/kexin type 9 inhibitors (PCSK9i) as adjunctive treatment in ICAS remains unclear.

**Methods:** We conducted a prospective, randomized, open-label, blinded-endpoint trial involving 62 patients with symptomatic ICAS. Participants were randomized 1:2 to receive either a PCSK9i (alirocumab 75 mg every two weeks) plus high-intensity statin or high-intensity statin alone for six months. The primary outcome was the change in intracranial artery stenosis measured by high-resolution vessel-wall magnetic resonance imaging. Secondary outcomes included plaque enhancement volume, low-density lipoprotein cholesterol (LDL-C) target achievement (<55 mg/dL), recurrent stroke events, modified Rankin Scale, and safety assessments.

**Results:** Among 62 participants (median age, 66 years; 68% men), 60 completed the study per protocol. At 6 months, median stenosis reduction was greater in the PCSK9i group (7.1%; 95% CI, 3.6–12.8%) than in controls (– 1.2%; 95% CI, −4.9–4.5%) (p < 0.01). Both groups showed significant reduction in plaque enhancement volume, but between-group difference was not significant (4.3 vs 4.0 mm^3^; p = 0.28). LDL-C <55 mg/dL was achieved in 85% of the PCSK9i group vs 13% of controls (p < 0.01). Recurrent stroke occurred in 5% vs 13% of patients (p = 0.39). No serious adverse events were reported in either group.

**Conclusion:** In patients with symptomatic ICAS, adjunctive PCSK9i therapy significantly reduced intracranial stenosis and improved LDL-C control over 6 months. While both treatment strategies reduced plaque enhancement, PCSK9i provided additional benefit in stenosis regression. Larger and longer-term studies are warranted to confirm these findings and clarify optimal LDL-C targets for ICAS management.

**Trial Registration:** ClinicalTrials.gov Identifier: NCT05001984

## 1 Introduction

Intracranial atherosclerotic stenosis (ICAS) is a major cause of ischemic stroke worldwide, particularly among Asian populations, ^1^ and is associated with a high risk of recurrent events despite standard secondary prevention. ^2,3^ Mechanisms of stroke recurrence in ICAS include hypoperfusion, artery-to-artery embolism, and branch atheromatous disease, and existing therapies—namely dual antiplatelet therapy and high-intensity statins—are often insufficient to fully mitigate this risk. ^4,5^

Intensive lipid lowering has been established as an effective strategy to reduce vascular events in atherosclerotic disease. The Stroke Prevention by Aggressive Reduction in Cholesterol Levels (SPARCL) trial demonstrated that high-intensity statin significantly reduces stroke and cardiovascular risk in patients with prior ischemic events, ^6,7^ while the Treat Stroke to Target (TST) trial showed an low-density lipoprotein cholesterol (LDL-C) target <70 mg/dL than that of 90-110mg/dL had a lower risk of subsequent cardiovascular events in patients with previous acute ischemic stroke or transient ischemic attack (TIA) with evidence of atherosclerosis. ^8^ High-intensity statin treatment and an LDL-C target <70 mg/dL is now recommended in patients with symptomatic ICAS. ^9-11^

Proprotein convertase subtilisin/kexin type 9 inhibitors (PCSK9i), such as alirocumab and evolocumab, have shown substantial benefit in reducing LDL-C and major cardiovascular events in high-risk populations. ^12-14^ Although their effect on coronary and carotid plaque burden has been validated, their impact on intracranial atherosclerosis remains less well defined. ^15^ Observational and imaging studies suggest that statin treatment may contribute to plaque regression and improved vessel stability in ICAS, potentially enhancing outcomes when used in conjunction with statins. ^16,17^

We hypothesized that the addition of PCSK9i to high-intensity statin therapy would promote intracranial plaque stabilization and regression in patients with symptomatic ICAS. To test this, we conducted the TOPICAL-MRI study (Trial of PCSK9 Inhibition in Patients with Acute Stroke and Symptomatic Intracranial Atherosclerosis) utilizing vessel-wall MRI to assess changes in intracranial plaque and clinical outcomes over a six-month treatment period. ^18^

## 2 Methods

### 2.1 Study Design and Participants

This prospective, open-label, blinded-endpoint trial was conducted at a tertiary medical center. ^18^ Patients with acute ischemic stroke or TIA within the past 7 days were enrolled. All participants had symptomatic ICAS (30%–99%) involving the intracranial internal carotid artery (ICA), middle cerebral artery (MCA) M1 segment, or basilar artery (BA)—confirmed by high-resolution vessel-wall MRI. All patients enrolled in this trial were classified as having large artery atherosclerosis based on clinical presentation and imaging findings, with other stroke etiologies (e.g., cardioembolism, dissection, vasculitis) excluded. ^19^ Stroke mechanisms associated with ICAS included: (1) branch atheromatous disease, (2) artery-to-artery embolism, (3) hypoperfusion or impaired embolic clearance distal to a stenosis, and (4) in situ thrombo-occlusion beyond a stenotic segment. ^4,20^ A baseline LDL-C ≥100 mg/dL or ≥70 mg/dL with ongoing statin therapy was required.

The study protocol was approved by the Institutional Review Board of Chang Gung Memorial Hospital (approval no. 202002482A3) and was registered on ClinicalTrials.gov (NCT05001984).

### 2.2 Randomization and Interventions

Participants were randomized in a 1:2 ratio to receive either: (1) a PCSK9i (alirocumab 75 mg subcutaneous every two weeks) plus high-intensity statin (rosuvastatin 20 mg or atorvastatin 40-80 mg daily), or (2) high-intensity statin alone. All patients received standardized stroke care, including dual antiplatelet therapy for 90 days, management of vascular risk factors, and structured lifestyle education. Participants underwent clinical and laboratory evaluations at baseline, 2 weeks, 3 months, and 6 months. Follow-up imaging with the same MRI protocol was performed at 6 months. LDL-C levels were monitored, and ezetimibe was permitted to achieve LDL-C <70 mg/dL, per guideline.

### 2.3 Study Outcomes

The primary endpoint was the changes in intracranial stenosis (%) of the target intracranial artery from baseline to 6 months on high-resolution MRI.

Secondary outcomes included: (1) change in plaque enhancement volume, (2) proportion achieving LDL-C <55 mg/dL at 6 months, (3) recurrent ischemic stroke or TIA in the same territory, (4) modified Rankin Scale (mRS) at 3 and 6 months, and (5) adverse events including hepatotoxicity, myopathy, injection-site reactions, and serious events.

### 2.4 MRI protocol and Image Analysis

MRI was performed using a 3.0 Tesla Siemens Verio system with a 32-channel head coil. Sequences included axial diffusion-weighted imaging (DWI), axial T1- and T2-weighted imaging, three-dimensional time-of-flight (TOF) magnetic resonance angiography (MRA), and dynamic susceptibility contrast perfusion imaging. High-resolution vessel-wall imaging was acquired using black-blood sequences with a spatial pre-saturation. Axial T2-weighted images were acquired with the following parameters: TR/TE = 3200/323 ms, echo train length = 185, slice thickness = 6 mm, flip angle = 180°, matrix size = 320 × 266, and field of view (FOV) = 149.6 mm. Pre- and post-contrast axial T1 fluid-attenuated inversion recovery images were obtained with TR/TE = 700/22 ms, echo train length = 55, slice thickness = 6 mm, flip angle = 180°, matrix = 320 × 320, and FOV = 190 mm. A gadolinium-based contrast agent was administered at a dose of 0.1 mmol/kg for post-contrast imaging. Sagittal views of T1-weighted, T2-weighted, and post-contrast T1-weighted images were reconstructed from the corresponding axial acquisitions, with the reconstruction plane oriented perpendicular to the long axis of the vessel at the site of the culprit stenosis to optimize visualization of the vessel wall and plaque characteristics.

A plaque was defined as wall thickening relative to adjacent or contralateral normal vessel segments. A culprit plaque was defined as either solitary or the most stenotic in the vascular territory corresponding to the infarct. Measurements were obtained by the cross-sectional of the culprit plaque. Vessel and lumen areas were manually traced and the wall area was calculated as the difference between vessel and lumen areas (Figure 1). Degree of stenosis was calculated as:

**Figure 1.**
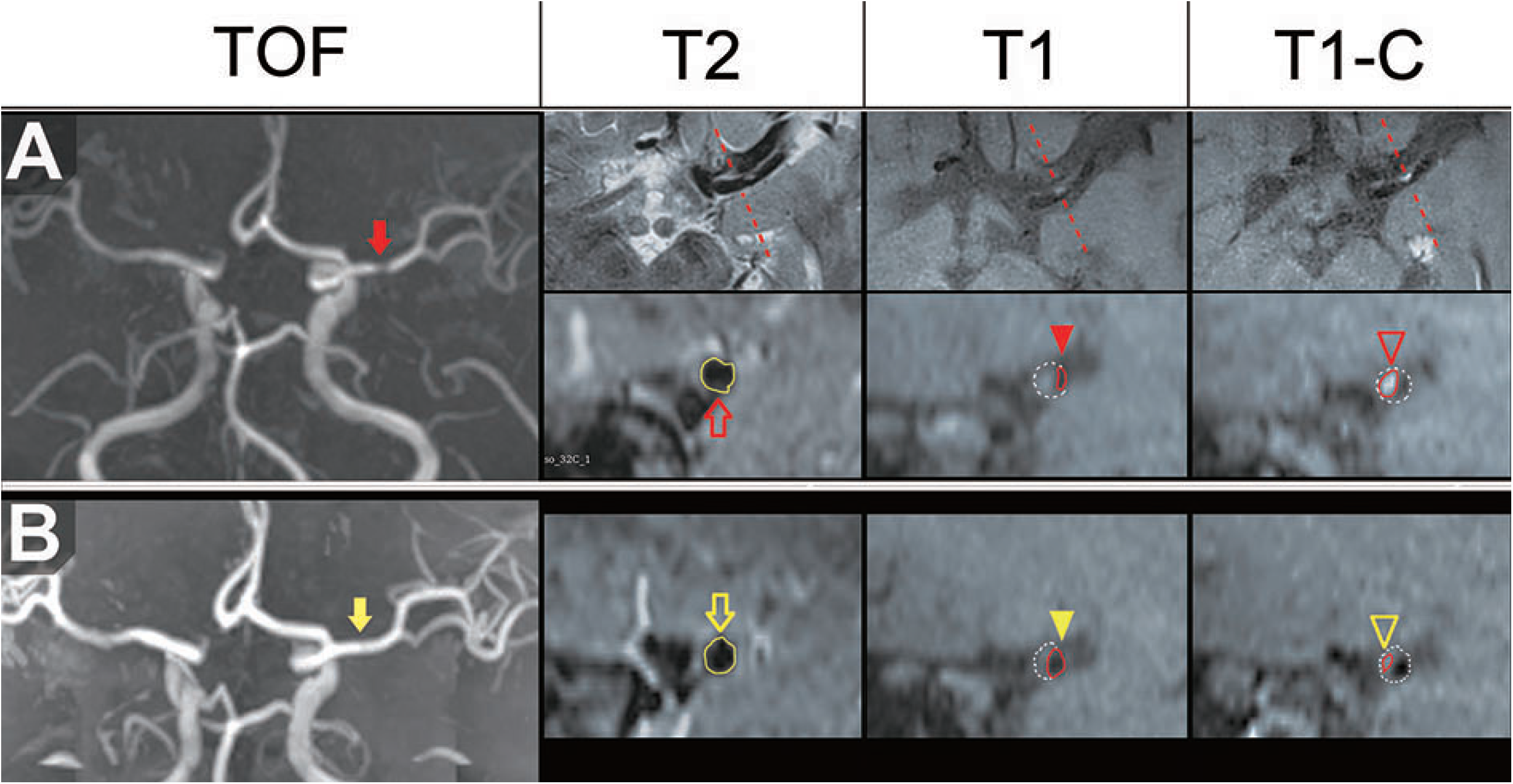
Illustration of high-resolution MRI measurements for intracranial atherosclerotic stenosis before and after treatment. (A) Baseline imaging of the maximal stenotic lesion. The site of maximal stenosis was identified at the left middle cerebral artery (MCA) on 3D time-of-flight (TOF) angiography (red arrow). Corresponding axial T2-weighted, T1-weighted, and T1-weighted contrast-enhanced images illustrate vessel-wall morphology. The vessel area (yellow outline, red hollow arrow) was measured at the proximal normal MCA on the T2-weighted image, while the lumen area (red circle, red solid arrowhead) and plaque enhancement area (red circle, red hollow arrowhead) were semi-automatically measured at the site of maximal stenosis. (B) Follow-up imaging at 6 months shows a reduction in stenosis severity and plaque enhancement. Follow-up TOF angiography (yellow arrow) demonstrates improved lumen diameter. Corresponding axial MRI sequences show the vessel area (yellow outline, yellow hollow arrow, T2-weighted image), increased lumen area (red circle, yellow solid arrowhead, T1-weighted image), and reduced plaque enhancement volume (red circle, yellow hollow arrowhead, T1-weighted contrast-enhanced image). All measurements were performed on sagittal images perpendicular to the long axis of the vessel at the site of maximal stenosis.

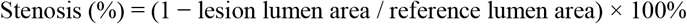

Plaque enhancement volume was quantified by comparing pre- and post-contrast T1-weighted images. Two independent, blinded readers performed all measurements; discrepancies were resolved by consensus.

### 2.5 Sample Size

The sample size was calculated using a two-sided t-test to detect an effect size of 0.7 between groups, assuming 80% power and α = 0.05. This moderate-to-large difference was based on prior ICAS data showing plaque regression with statins. ^16^ Although vessel-wall imaging data were unavailable during study design, the estimate was considered biologically plausible. A total of 60 patients (20 intervention, 40 control) was required.

### 2.6 Statistical analysis

All statistical analyses were performed using SPSS version 27 (IBM Corp., Armonk, NY). Normality of continuous variables was assessed via the Kolmogorov–Smirnov test. Non-normally distributed data were summarized as medians with interquartile ranges and compared using the Mann– Whitney U test. Categorical variables were analyzed with Chi-square or Fisher’s exact tests.

The primary endpoint, percentage change in intracranial stenosis, was analyzed using a rank-based analysis of covariance adjusting for baseline stenosis, age, and baseline NIHSS score. Binary secondary outcomes were analyzed using generalized linear models with a Poisson distribution, log link function, and robust standard errors to estimate adjusted risk ratios.

Within-group laboratory changes used the Wilcoxon signed-rank test. Inter-rater reliability for stenosis measurement was assessed by average-measures intraclass correlation coefficient (ICC) using a two-way random effects model with absolute agreement definition. As the final stenosis values were averaged across two independent raters, the average-measures ICC was reported to reflect the reliability of these aggregated scores. Spearman’s rank correlation coefficient was used to assess associations between LDL-C levels (or change) and stenosis reduction. Scatterplots with overlaid linear regression lines and 95% confidence intervals (CI) were used for visual presentation of the trends. All statistical tests were two-tailed, and a p-value < 0.05 was considered statistically significant.

## 3 Results

### 3.1 Patient Enrollment and Baseline Characteristics

Between July, 2021, and October, 2024, 1859 patients were screened, of whom 1797 were excluded (Figure 2). A total of 62 patients met eligibility criteria and were enrolled in the intention-to-treat (ITT) population—20 in the PCSK9i group and 42 in the control group. Two patients in the control group did not undergo follow-up MRI, resulting in a per-protocol (PP) population of 60 patients (20 PCSK9i, 40 control).

**Figure 2.**
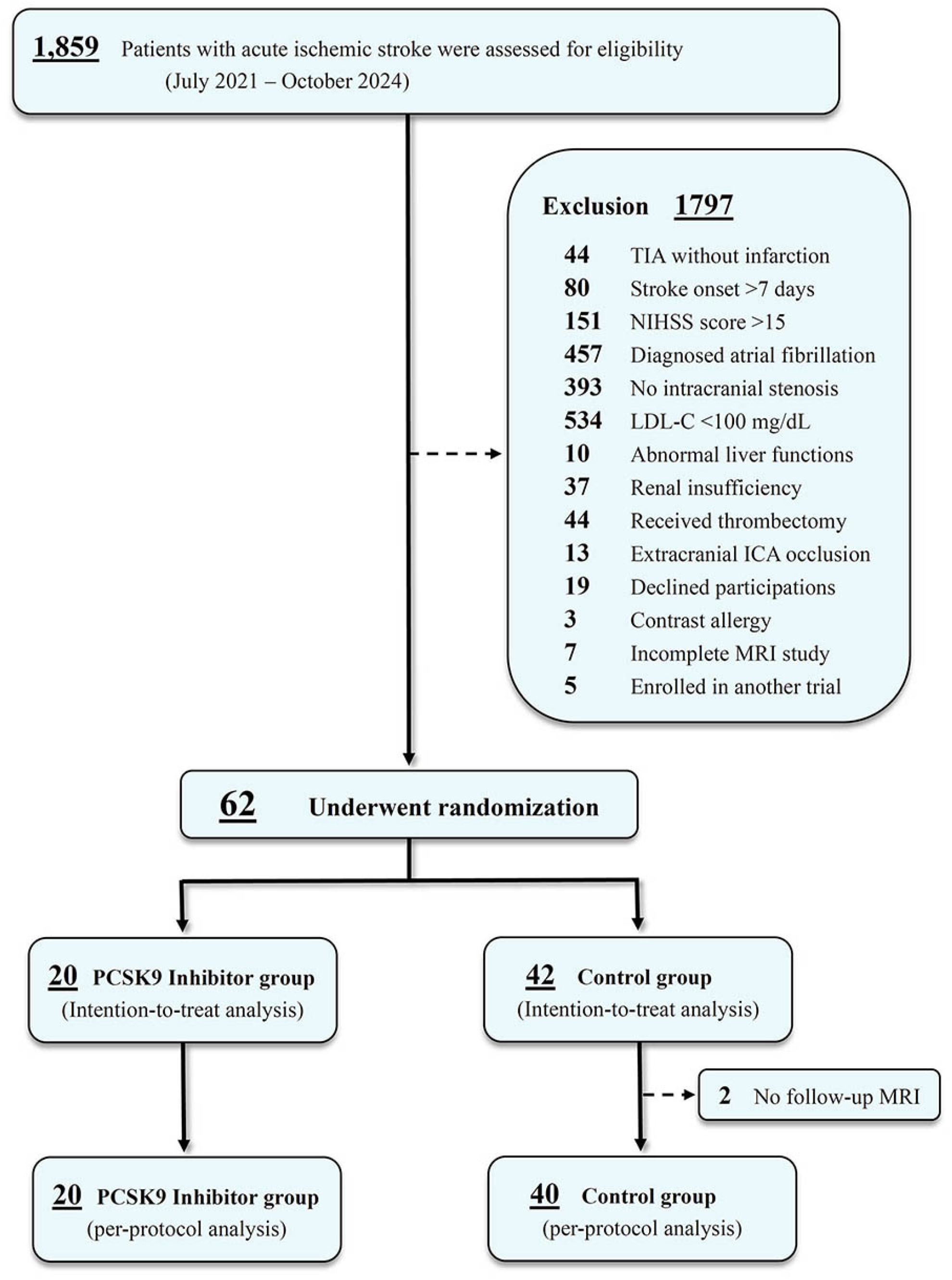
Study Flowchart of TOPICAL-MRI Trial. Abbreviations: ICA, internal carotid artery; LDL-C, low-density lipoprotein cholesterol; PCSK9i, proprotein convertase subtilisin/kexin type 9 inhibitor.

Baseline characteristics were well balanced between groups in the ITT population (Table 1), including age (median 66 years, IQR 52–72), vascular risk factors, baseline NIHSS score (median 3, IQR 1–5), time from stroke onset to randomization (median 4 days, IQR 3–6), and interval to follow-up MRI (median 184 days, IQR 180–193). Culprit plaques were located in the ICA in 5 patients (8%), the MCA in 33 patients (53%), and the BA in 24 patients (39%). The most common stroke mechanism was branch atheromatous disease (44%), followed by hypoperfusion or impaired embolic clearance (27%). Initial stenosis was greater in the PCSK9i group compared to the control group (85.6% vs. 77.5%).

**Table 1.** Baseline Characteristics of the Study Groups.

| **Characteristic** | **PCSK9i**<br>**(N = 20)** | **Control**<br>**(N = 42)** |
| --- | --- | --- |
| Age, median (IQR), years | 66 (52–72) | 69 (58–79) |
| Female, No. (%) | 6 (30) | 8 (19) |
| **Past medical history, No. (%)** |  |  |
| Hypertension | 18 (90) | 35 (83) |
| Diabetes mellitus | 13 (65) | 22 (52) |
| Hypercholesterolemia | 15 (75) | 33 (79) |
| Coronary artery disease | 1 (5) | 0 (0) |
| Congestive heart failure | 1 (5) | 1 (2.4) |
| Prior stroke | 3 (15) | 8 (19) |
| Current smoking | 8 (40) | 16 (38) |
| Baseline antiplatelet use | 4 (20) | 6 (14) |
| Baseline statin use | 5 (25) | 10 (24) |
| **Vital signs and lab data, median (IQR)** |  |  |
| Systolic BP, mmHg | 167 (144–188) | 159 (148–196) |
| Diastolic BP, mmHg | 101 (93–116) | 103 (86–122) |
| Total cholesterol, mg/dL | 200 (171–236) | 197 (171–219) |
| LDL-C, mg/dL | 142 (106–167) | 131 (112–152) |
| Triglyceride, mg/dL | 142 (100–191) | 126 (103–161) |
| Glycohemoglobin, % | 6.3 (5.7–7.3) | 6.3 (5.7–7.2) |
| **Stroke information** |  |  |
| Onset to randomization, median (IQR), days | 5 (3–6) | 4 (3–6) |
| NIHSS score on arrival, median (IQR) | 3 (1–5) | 3 (1–5) |
| Statin types, No. (%) |  |  |
| Atorvastatin 40 mg | 8 (40) | 12 (29) |
| Rosuvastatin 20 mg | 12 (60) | 30 (71) |
| Vascular territory of infarction, No. (%) |  |  |
| Middle cerebral artery | 12 (60) | 21 (50) |
| Internal carotid artery | 3 (15) | 2 (5) |
| Basilar artery | 5 (25) | 19 (45) |
| Stenosis, median (IQR), % | 85.6 (73.4–88.5) | 77.5 (65.3–87.8) |
| Infarct mechanisms, No. (%) |  |  |
| Artery-to-artery embolization | 1 (5) | 6 (14) |
| Branch atheromatous disease | 7 (35) | 20 (48) |
| Hypoperfusion | 8 (40) | 9 (21) |
| In situ thrombo-occlusion | 0 (0) | 3 (7) |
| Mixed | 4 (20) | 4 (10) |
| Time interval of follow-up imaging, median (IQR), days | 183 (181–192) | 183 (179–195) |

Abbreviations: BP, blood pressure; HDL-C, high-density lipoprotein cholesterol; LDL-C, low-density lipoprotein cholesterol; NIHSS, National Institutes of Health Stroke Scale; PCSK9i, proprotein convertase subtilisin/kexin type 9 inhibitor.

### 3.2 Primary Outcome

To ensure measurement consistency, inter-rater reliability was assessed. The intraclass correlation coefficient (ICC) for pre-treatment stenosis measurements was 0.72 (95% CI, 0.49–0.84), and for post-treatment stenosis was 0.70 (95% CI, 0.49–0.82), indicating moderate to good agreement between raters. The primary outcome—change in intracranial stenosis—showed a significant between-group difference at 6 months (Table 2). The PCSK9i group had a median stenosis reduction of 7.1% (95% CI, 3.6–12.8%), while the control group had no meaningful change (−1.2%, 95% CI, −4.9–4.5%) (p < 0.01, adjusted for baseline stenosis).

**Table 2.** Primary and Secondary Efficacy Outcomes, Per-Protocol Analysis.

| End point | PCSK9i<br>(N = 20) | Control<br>(N = 40) | Relative Risk<br>(95% CI) | p-value |
| --- | --- | --- | --- | --- |
| **Primary Efficacy Outcome** |  |  |  |  |
| Stenosis degree reduction, median (IQR), % | 7.1 (3.6–12.8) | -1.2 (-4.9–4.5) |  | <0.01* |
| **Secondary Efficacy Outcomes** |  |  |  |  |
| Enhancement volume reduction, median (IQR), mm<sup>3</sup> | 4.3 (0.5–15.5) | 4.0 (0–8.4) |  | 0.28* |
| Major cardiovascular events within 6 months, No. (%) | 2 (10) | 5 (13) | 0.73 (0.14–3.85) † | 0.71 |
| Recurrent stroke within 6 months, No. (%) | 1 (5) | 5 (13) | 0.39 (0.04–3.41) † | 0.39 |
| Achieved LDL-C goals (<70 mg/dL), No. (%) | 19 (95) | 24 (60) | 1.56 (0.84–2.88) † | 0.16 |
| Achieved LDL-C goals (<55 mg/dL), No. (%) | 17 (85) | 5 (13) | 6.51 (2.37–17.9) † | <.01 |
| Good outcomes (mRS≤2) at 6 months, No. (%) ‡ | 16 (80) | 35 (88) | 0.91 (0.50–1.66) † | 0.75 |

\* Adjusted for baseline stenosis/enhancement volume, age, and NIHSS score using rank-based analysis of covariance.
† Risk ratio for PCSK9 inhibitor therapy were adjusted for age, initial NIHSS score and baseline LDL-C level, but not for multiple comparisons.
‡ Scores on the mRS of functional disability range from 0 (no symptoms) to 6 (death).
Abbreviations: LDL-C, Low-density lipoprotein cholesterol; mRS: modified Rankin Scale; NIHSS, National Institutes of Health Stroke Scale; PCSK9i, proprotein convertase subtilisin/kexin type 9 inhibitor.

### 3.3 Secondary Outcomes and Safety Events

Secondary outcomes showed that the reduction in plaque enhancement volume was greater in the PCSK9i group (mean change: 4.3 mm^3^) compared to the control group (4.1 mm^3^); however, this difference was not statistically significant (p = 0.28; Table 2). A significantly higher proportion of patients in the PCSK9i group achieved the target LDL-C level of <55 mg/dL (85% vs. 13%; adjusted RR 6.51, 95% CI 2.37–17.9). In contrast, there were no significant between-group differences in the rates of major cardiovascular events (10% vs. 13%, p = 0.71), recurrent stroke (5% vs. 13%, p = 0.39), or favorable functional outcomes (mRS ≤2: 80% vs. 88%, p = 0.75). No deaths, major bleeding, or severe adverse events occurred. Liver function abnormalities were observed in 2 patients (10%) in the PCSK9i group and 6 (15%) in the control group (Supplement 1, Table S1).

### 3.4 Laboratory and Imaging Analysis

Lipid profile improvements were significantly greater in the PCSK9i group. At 6 months, LDL-C decreased from 142 mg/dL to 25 mg/dL in the PCSK9i group, and from 131 mg/dL to 69 mg/dL in the control group (25 vs 69 mg/dL, p < 0.01). Total cholesterol and LDL-C levels decreased significantly in both groups, while high-density lipoprotein cholesterol (HDL-C) increased (Supplement 1, Table S2). Triglyceride levels also decreased, with significance reached only in the control group. There were no significant between-group differences in aspartate transaminase, alanine aminotransferase, creatine phosphokinase or glycohemoglobin.

Stenosis was significantly reduced after 6 months in the PCSK9i group (85.6% to 75.7%, p < 0.01), while no significant change was observed in the control group (77.5% to 75.8%, p = 0.63). Plaque enhancement volume decreased significantly in both groups: from 25.1 to 10.1 mm^3^ in the PCSK9i group (p = 0.01), and from 11.3 to 6.6 mm^3^ in the control group (p < 0.01). As shown in Supplement 1 Figure S1, LDL-C levels were significantly lower in the PCSK9i group compared to controls at both 3 and 6 months.

### 3.5 Correlation Between LDL-C and Stenosis Improvement

A significant inverse correlation was observed between LDL-C levels at 3 months and stenosis reduction at 6 months (Spearman’s ρ = –0.47, p < 0.01), indicating that lower LDL-C levels were associated with greater stenosis improvement (Figure 3). Additionally, absolute LDL-C reduction (baseline to 3 months) was positively correlated with stenosis reduction (Spearman’s ρ = 0.45, p < 0.01), further supporting the impact of lipid lowering on plaque regression.

**Figure 3.**
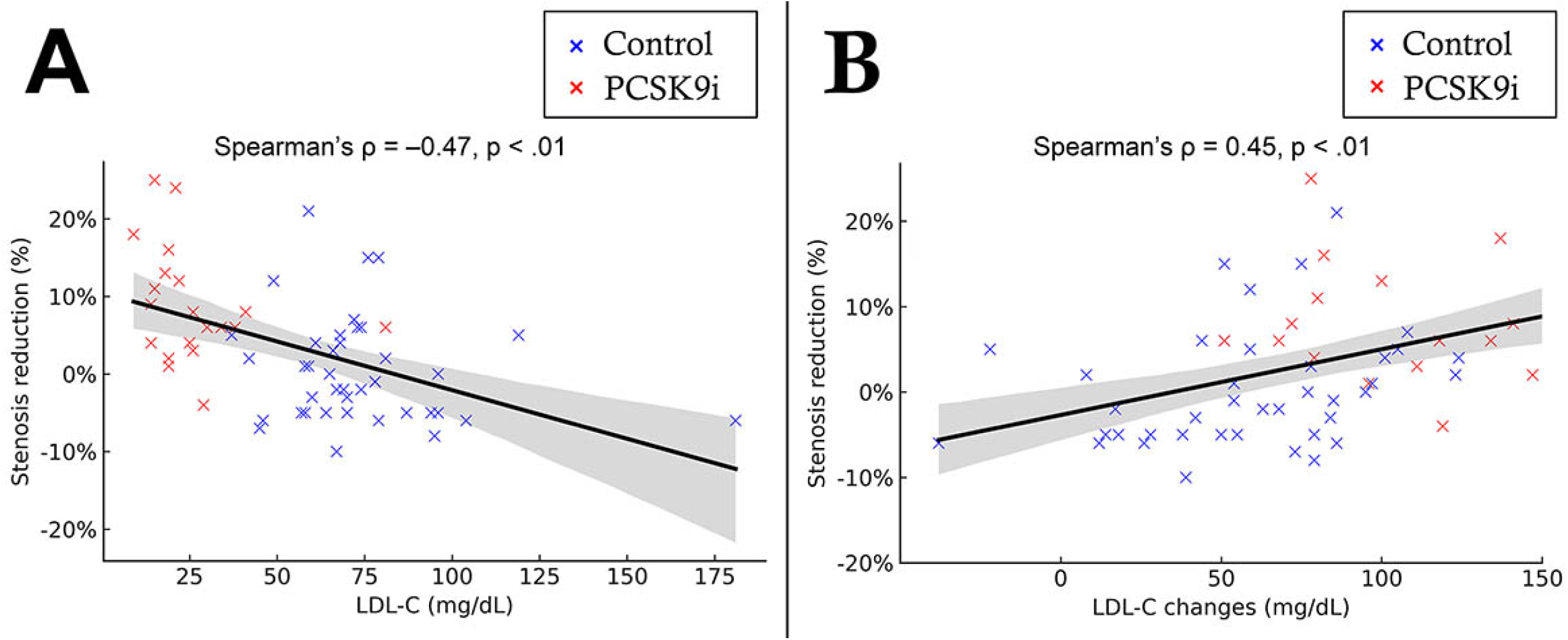
Association Between LDL-C Levels and Intracranial Stenosis Reduction at 6 Months. (A) Correlation between post-treatment low-density lipoprotein cholesterol (LDL-C) levels at 3 months and percent stenosis reduction. (B) Correlation between absolute LDL-C reduction and percent stenosis reduction. Each dot represents an individual patient, with red indicating the intervention group (proprotein convertase subtilisin/kexin type 9 inhibitor [PCSK9i] plus statin) and blue indicating the control group (statin alone). Linear regression lines are displayed for both groups, with shaded areas indicating the 95% confidence intervals of the regression estimates. A significant inverse correlation was observed between post-treatment LDL-C levels and stenosis reduction (Spearman’s ρ = –0.47, p < 0.01), suggesting that lower LDL-C is associated with greater plaque regression. A significant positive correlation was found between absolute LDL-C reduction and stenosis improvement (Spearman’s ρ = 0.45, p < 0.01), indicating that greater LDL-C reduction is associated with more pronounced vascular benefit.

## 4 Discussion

This randomized controlled trial provides compelling evidence that adjunctive therapy with a PCSK9i significantly reduces intracranial stenosis and LDL-C levels in patients with symptomatic ICAS relative to standard lipid-lowering therapy alone. These findings align with prior studies, underscoring that PCSK9i achieves substantial LDL-C reductions and may promote plaque stabilization, thereby reducing the risk of early stroke recurrence.

Compelling evidence from recent coronary imaging trials examining PCSK9i combined with statin therapy demonstrates a consistent, linear association between achieved LDL-C levels and favorable coronary plaque changes. ^21-23^ These include reductions in percent atheroma volume and increases in fibrous cap thickness, with beneficial effects observed at LDL-C levels below 20 mg/dL. ^21^ Our results are further corroborated by recent intracranial-specific studies. In a prospective single-arm study, Zeng et al. reported significant reductions in both stenosis severity and plaque enhancement after 24 weeks of combined high-intensity statin and evolocumab therapy. ^24^ Importantly, a strong correlation was observed between LDL-C reductions and plaque contrast volume, reinforcing the concept that PCSK9i promote intracranial plaque stabilization and vascular remodeling. Similarly, Wu et al. demonstrated that statins and PCSK9i significantly reduced plaque enhancement volume and stenosis degree in the observation study. ^25^ These data reinforce the potential of aggressive lipid-lowering to stabilize intracranial plaques and reduce recurrent cerebrovascular events.

It is important to contextualize our findings within the current framework of lipid management guidelines. Current lipid guidelines recommend LDL-C <70 mg/dL for high-risk patients, yet do not specify thresholds for ICAS. ^10^ Notably, the 2022 European Stroke Organisation guidelines on treatment of patients with ICAS suggest that isolated symptomatic ICAS should be considered a very-high-risk condition—targeting LDL-C levels below 55 mg/dL. ^26^ Our study provides valuable preliminary evidence that such aggressive lipid targets may be particularly beneficial in this population and warrant further investigation.

Both treatment groups in our trial showed significant reductions in plaque enhancement volume; however, between-group differences were not significant. Plaque enhancement reflects neovascularization, inflammation, and endothelial dysfunction^27^—features linked to plaque instability and heightened stroke risk. ^28-30^ These findings suggest that statins alone may confer strong anti-inflammatory effects sufficient to reduce enhancement volume. ^15,17^ Prior research has shown that statins can stabilize plaque independent of LDL-C lowering. ^31^ Therefore, the similar reductions in enhancement volume between groups likely reflect the robust anti-inflammatory activity of statins, reinforcing their foundational role in ICAS treatment.

PCSK9 inhibitors add-on therapy further lowered LDL-C levels and reduced early recurrent stroke risk in patients with symptomatic ICAS, particularly within the first month poststroke. ^32^ However, our study did not detect significant differences in recurrent stroke or major cardiovascular events between groups, likely due to limited sample size and follow-up. Nevertheless, the safety profile of PCSK9i was reaffirmed, with low adverse event rates and no serious safety concerns, consistent with prior cardiovascular outcome trials. Trials such as SAMMPRIS and CASSISS underscore the challenges in managing ICAS, marked by high stroke recurrence and uncertain benefit from invasive interventions like stenting. ^11,33,34^ These findings support the value of optimized medical therapy, including dual antiplatelet and intensive lipid-lowering regimens. Our study adds evidence for PCSK9i as adjunctive therapy to statins in comprehensive ICAS care. Larger trials with extended follow-up are needed to assess long-term outcomes and define optimal treatment targets.

### Limitations

Several limitations warrant careful consideration. First, the small sample size and six-month follow-up limit power to detect clinical outcome differences. Although we demonstrated significant imaging changes, larger studies with longer duration are required to confirm the clinical significance of plaque regression. Prior ICAS trials (e.g., SAMMPRIS, CASSISS) emphasize the importance of sufficient sample sizes and extended follow-up to capture rare or delayed events. ^11,33,34^

Second, the open-label design may introduce bias. Although imaging assessors were blinded, the lack of patient and clinician blinding could affect medication adherence or management. Future double-blind, placebo-controlled studies would mitigate such bias and strengthen evidence of efficacy. Third, although high-resolution vessel-wall MRI is a well-established method for evaluating intracranial plaque morphology, it has inherent limitations. Vessel-wall imaging lacks direct histopathological validation, which means MRI findings such as enhancement or plaque regression cannot be conclusively correlated with histological processes like neovascularization or inflammation. ^30,35^ Pathological validations through multimodal imaging and histology would significantly enhance the interpretation of these MRI-based findings. ^36^

Furthermore, generalizability may be limited due to the trial’s selective criteria. Patients with renal/hepatic dysfunction, prior intolerance to statins or PCSK9i, or MRI contraindications were excluded, potentially restricting applicability. Larger ongoing studies like PISTIAS, which employ broader eligibility criteria, will offer further insights into the generalizability of PCSK9i therapy for ICAS. ^37^ Finally, the optimal LDL-C target in ICAS patients remains an area of uncertainty. While our findings support LDL-C lowering below 55 mg/dL, current guidelines still recommend <70 mg/dL for high-risk cerebrovascular patients. ^38^ Future trials designed specifically to explore various LDL-C targets in ICAS could provide critical data to guide clinical decision-making and guideline updates.

In summary, while this trial presents promising data on PCSK9i therapy for symptomatic ICAS, these limitations highlight important areas for further research. Larger, rigorously designed randomized trials incorporating multimodal imaging, extended follow-up, and broader populations are essential to validate and expand upon our results.

## Conclusion

The addition of PCSK9i to high-intensity statin therapy significantly reduced intracranial stenosis and improved LDL-C goal achievement in patients with symptomatic ICAS, with encouraging signs of plaque stabilization. While both regimens appeared effective in reducing plaque inflammation, PCSK9i conferred added benefit in stenosis regression. These findings support the potential of PCSK9i in ICAS management and highlight the need for large-scale trials to confirm long-term efficacy and clarify LDL-C targets in this high-risk population.

## Data Availability

Data supporting the findings of this study are available from the corresponding author upon reasonable request.

## Nonstandard Abbreviations and Acronyms

BA: Basilar Artery
CI: Confidence Interval
DWI: Diffusion-Weighted Imaging
FOV: Field of View
HDL-C: High-Density Lipoprotein Cholesterol
ICA: Internal Carotid Artery
ICAS: Intracranial Atherosclerotic Stenosis
ICC: Intraclass Correlation Coefficient
ITT: Intention-to-Treat
LDL-C: Low-Density Lipoprotein Cholesterol
MCA: Middle Cerebral Artery
MRI: Magnetic Resonance Imaging
MRA: Magnetic Resonance Angiography
mRS: Modified Rankin Scale
NIHSS: National Institutes of Health Stroke Scale
PCSK9i: Proprotein Convertase Subtilisin/Kexin Type 9 Inhibitor
PP: Per-Protocol
SPARCL: Stroke Prevention by Aggressive Reduction in Cholesterol Levels
TIA: Transient Ischemic Attack
TOF: Time-of-Flight
TST: Treat Stroke to Target

## Ethics approval and consent to participate

The study protocol was approved by the Institutional Review Board of Chang Gung Memorial Hospital (approval no. 202002482A3) and was registered on ClinicalTrials.gov (NCT05001984). Prior to participation, all enrolled individuals provided written informed consent.

## Consent for publication

Not applicable.

## Acknowledgments

We thank Yi-Chen Kuo for assisting us with this study.

## Sources of Funding

The TOPICAL-MRI trial was funded by National Science and Technology Council (MOST 110-2314-B-182-056, 111-2314-B-182 −055, 112-2314-B-182-041, 113-2314-B-182 −061) and Chang Gung Memorial Hospital (CMRPG6P0231).

## Competing interests

None declared.

## Disclosures

none

## Notes

### Competing Interest Statement

The authors have declared no competing interest.

### Clinical Trial

ClinicalTrials.gov Identifier: NCT05001984

### Author Declarations

The study protocol was approved by the Institutional Review Board of Chang Gung Memorial Hospital (approval no. 202002482A3)

